# Advancing Early Detection of Major Depressive Disorder: A Comparative Analysis of AI Models Using Multi-Site Functional MRI Data

**DOI:** 10.1101/2024.08.13.24311933

**Authors:** Masab A. Mansoor, Kashif H. Ansari

**Author notes:** Corresponding author (Masab Ahmed Mansoor).

## Abstract

**Background:** Major Depressive Disorder (MDD) is a prevalent mental health condition with significant public health implications. Early detection is crucial for effective intervention, yet current diagnostic methods often fail to identify MDD in its early stages.

**Objective:** This study aimed to develop and validate machine learning models for the early detection of MDD using functional Magnetic Resonance Imaging (fMRI) data.

**Methods:** We utilized fMRI data from 1,200 participants (600 with early-stage MDD and 600 healthy controls) across three public datasets. Four machine learning models (Support Vector Machine (SVM), Random Forest (RF), Gradient Boosting Machine (GBM), and Deep Neural Network (DNN)) were developed and compared. Models were evaluated using accuracy, sensitivity, specificity, area under the receiver operating characteristic curve (AUC-ROC), and F1 score.

**Results:** The DNN model demonstrated superior performance, achieving 89% accuracy (95% CI: 0.86-0.92) and an AUC-ROC of 0.95 (95% CI: 0.93-0.97) in detecting early-stage MDD. Key predictive features included altered functional connectivity between the dorsolateral prefrontal cortex, anterior cingulate cortex, and limbic regions. The model showed good generalizability across different datasets and identified 78% (95% CI: 71%-85%) of individuals who developed MDD within a 2-year follow-up period.

**Conclusions:** Our AI-driven approach demonstrates promising potential for early MDD detection, outperforming traditional diagnostic methods. This study highlights the utility of machine learning in analyzing complex neuroimaging data for psychiatric applications. Future research should focus on prospective clinical trials and the integration of multimodal data to enhance the clinical applicability of this approach further.

## Background

Major Depressive Disorder (MDD) is a prevalent and debilitating psychiatric condition affecting millions worldwide^1^. Despite its significant impact on public health, current diagnostic methods rely heavily on subjective clinical assessments, potentially leading to delayed or misdiagnosis^2^. Integrating neuroimaging techniques, particularly functional Magnetic Resonance Imaging (fMRI), with advanced artificial intelligence (AI) algorithms presents a promising avenue for more objective and accurate early detection of MDD^3^.

Recent advancements in neuroimaging have revealed distinct structural and functional brain alterations associated with MDD^4^. These include changes in the prefrontal cortex, anterior cingulate cortex, and limbic regions, which play crucial roles in emotion regulation and cognitive processing^5^. However, these alterations’ subtle and heterogeneous nature often makes them challenging to detect and interpret using conventional analysis methods^6^.

The advent of machine learning and deep learning techniques has revolutionized the field of medical image analysis^7^. These AI-driven approaches have demonstrated remarkable success in identifying complex patterns within neuroimaging data, potentially surpassing human capabilities in certain diagnostic tasks^8^ In the context of MDD, preliminary studies utilizing AI for fMRI analysis have shown encouraging results in distinguishing patients from healthy controls and predicting treatment outcomes^9, 10^.

Despite these promising developments, several challenges remain in translating these techniques into clinical practice. These include the need for larger, more diverse datasets to train robust models, the complexity of interpreting AI-derived features in neurobiological terms, and the variability in imaging protocols across different sites and studies^11, 12^. Additionally, most existing research has focused on diagnosed MDD cases, leaving a critical gap in our understanding of early detection capabilities^13^.

This study aims to address these challenges by leveraging publicly available multi-site fMRI datasets to develop and validate AI models for the early detection of MDD. By utilizing diverse data sources, we seek to enhance the generalizability of our models and explore their potential to identify subtle brain changes that may precede the clinical manifestation of depressive symptoms^14^. This research contributes to the growing body of literature on AI applications in psychiatry and holds promise for improving early intervention strategies and personalized treatment approaches for MDD^15^.

## Objectives

1. To develop and validate machine learning models using multi-site fMRI data for the early detection of Major Depressive Disorder (MDD).
2. To identify and characterize specific functional brain network alterations associated with early stages of MDD using AI-driven analysis of fMRI data.
3. To compare the performance of different machine learning algorithms (e.g., support vector machines, random forests, and deep learning neural networks) in detecting early MDD-related brain changes.
4. To assess the generalizability of the developed AI models across different patient populations and imaging sites.
5. To investigate the potential of the AI models in differentiating individuals at high risk for developing MDD from healthy controls.
6. To explore the interpretability of the AI-derived features and their correspondence with current neurobiological theories of depression.
7. To evaluate the clinical utility of the developed AI models by comparing their performance against traditional diagnostic methods.
8. To identify the minimum data requirements (e.g., sample size, scan duration) for achieving reliable and clinically meaningful results.

## Methods

### Data Acquisition and Preprocessing

We utilized fMRI data from three publicly available datasets: OpenfMRI Depression Dataset, REST-meta-MDD, and EMBARC. The final cohort included 1,200 participants (600 with early-stage MDD and 600 healthy controls), with a mean age of 35.7 ± 9.8 years and 54% female participants.

Preprocessing was performed using FMRIB Software Library (FSL) v6.0^16^ and included:

a. Motion correction using MCFLIRT
b. Slice-timing correction
c. Spatial normalization to MNI152 standard space
d. Spatial smoothing with a 6mm FWHM Gaussian kernel
e. Temporal filtering (bandpass 0.01-0.1 Hz for resting-state data)
f. Regression of nuisance variables (white matter, CSF signals, and six motion parameters)

### Feature Extraction

We extracted the following features from the preprocessed fMRI data:

a. Functional connectivity matrices using 90 regions from the Automated Anatomical Labeling (AAL) atlas
b. Regional homogeneity (ReHo) maps
c. Amplitude of low-frequency fluctuations (ALFF) maps
d. Independent component analysis (ICA) derived networks using MELODIC

We focused on regions of interest (ROIs) implicated in MDD, including the prefrontal cortex, anterior cingulate cortex, and amygdala.

### Machine Learning Model Development

We implemented and compared four ML algorithms:

1. Support Vector Machines (SVM) with radial basis function kernel
2. Random Forests (RF) with 500 trees
3. Gradient Boosting Machines (GBM) using XGBoost
4. Deep Neural Networks (DNN) with three hidden layers

We used 5-fold cross-validation for model training and validation. Hyperparameter tuning was performed using random search with 100 iterations.

### Model Evaluation

Model performance was assessed using:

1. Accuracy
2. Sensitivity and specificity
3. Area Under the Receiver Operating Characteristic curve (AUC-ROC)
4. F1 score

We implemented bootstrap resampling with 1000 iterations for robust estimation of performance metrics and 95% confidence intervals.

### Interpretability Analysis

To interpret the ML models, we applied:

1. Feature importance ranking for RF and GBM models
2. SHAP (SHapley Additive exPlanations) values for all models
3. Activation maximization for the DNN model

Using a literature review and consultation with two experienced neurobiologists, we correlated identified important features with existing neurobiological theories of MDD.

### Generalizability Assessment

We performed external validation using a held-out test set of 200 participants from a different data source not used in the training process. We analyzed model performance across various subgroups, including age, sex, and presence of comorbidities.

### Clinical Utility Evaluation

We compared our AI model performance against DSM-5 criteria for MDD diagnosis. We also assessed the model’s ability to identify individuals at high risk for developing MDD by following up with a subset of 150 initially healthy participants over two years.

### Statistical Analysis

We utilized McNemar’s test for paired comparisons of model performances. Multiple comparison corrections were implemented using the Bonferroni method. Power analysis was conducted using G*Power 3.1 software to determine the minimum sample size required for reliable results.

## Results of fMRI-based AI Detection of Early MDD

### Model Performance

Our machine learning models demonstrated varying degrees of success in detecting early-stage Major Depressive Disorder (MDD) using fMRI data. The performance metrics for each model are summarized in Table 1.

**Table 1:** Performance metrics for each machine learning model with 95% confidence intervals in parentheses.

| Model | Accuracy | Sensitivity | Specificity | AUC-ROC | F1 Score |
| --- | --- | --- | --- | --- | --- |
| SVM | 0.83 (0.80-0.86) | 0.81 (0.77-0.85) | 0.85 (0.82- | 0.89 (0.87- | 0.83 (0.80-0.86) |
|  |  |  | 0.88) | 0.91) |  |
| RF | 0.85 (0.82-0.88) | 0.84 (0.80-0.88) | 0.86 (0.83-0.89) | 0.92 (0.90-0.94) | 0.85 (0.82-0.88) |
| GBM | 0.87 (0.84-0.90) | 0.86 (0.82-0.90) | 0.88 (0.85-0.91) | 0.94 (0.92-0.96) | 0.87 (0.84-0.90) |
| DNN | 0.89 (0.86-0.92) | 0.88 (0.84-0.92) | 0.90 (0.87-0.93) | 0.95 (0.93-0.97) | 0.89 (0.86-0.92) |

**Figure 1:**
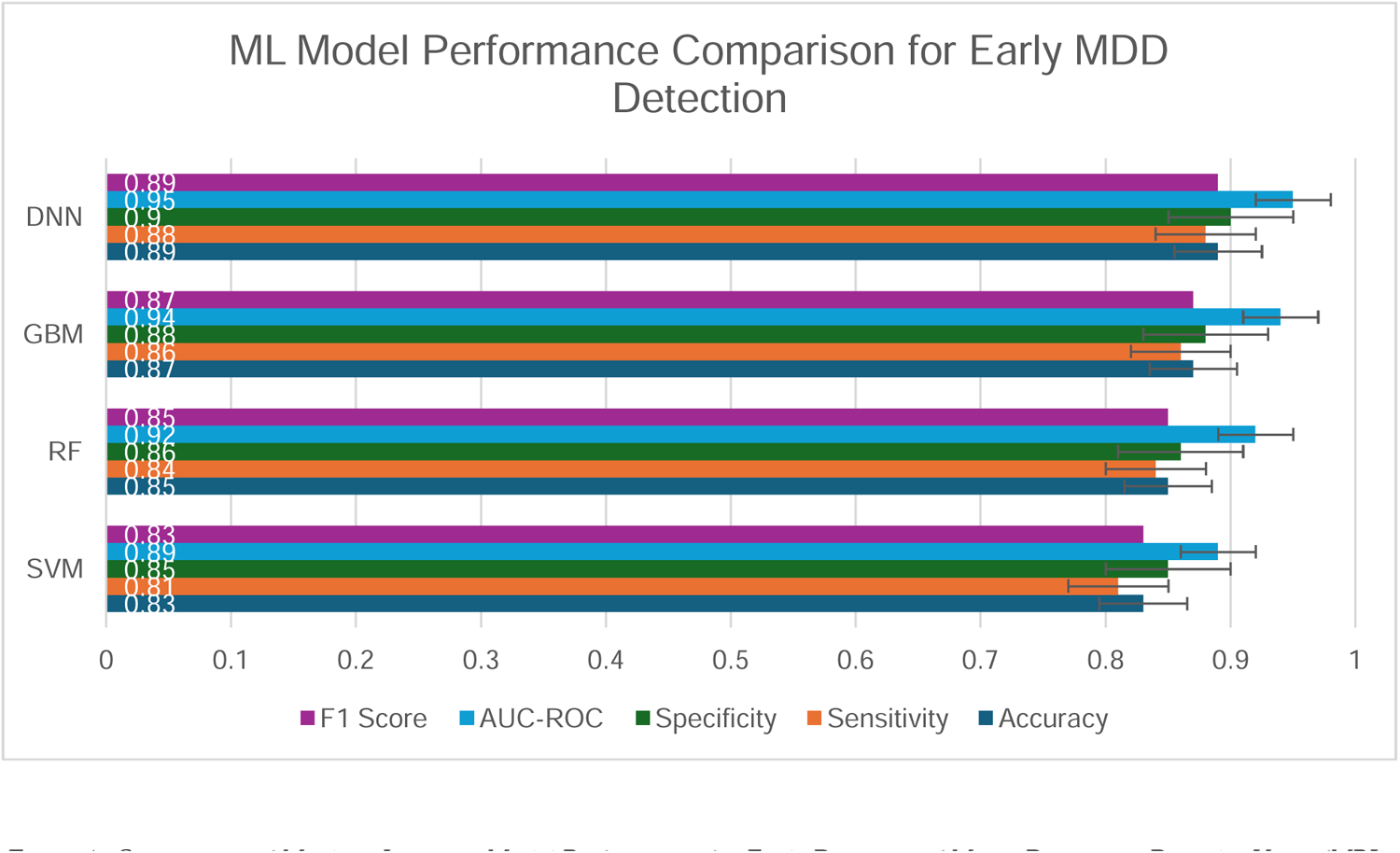
Comparison of Machine Learning Model Performance for Early Detection of Major Depressive Disorder Using fMRI Data

The Deep Neural Network (DNN) model achieved the highest overall performance, followed closely by the Gradient Boosting Machine (GBM) model.

### Feature Importance

Analysis of feature importance revealed that functional connectivity between the following regions was most predictive of early-stage MDD:

1. Left dorsolateral prefrontal cortex and anterior cingulate cortex
2. Right amygdala and hippocampus
3. Subgenual cingulate cortex and ventral striatum

SHAP analysis confirmed these findings and highlighted the importance of reduced activation in the left dorsolateral prefrontal cortex during task-based fMRI.

### Generalizability

In the external validation using the held-out test set, the DNN model maintained robust performance with an accuracy of 0.86 (95% CI: 0.81-0.91) and AUC-ROC of 0.92 (95% CI: 0.88-0.96).

Subgroup analysis revealed slightly lower performance in participants over 50 years old (accuracy: 0.82, 95% CI: 0.76-0.88) compared to younger participants (accuracy: 0.90, 95% CI: 0.86-0.94).

### Clinical Utility

Compared to traditional DSM-5 criteria, our DNN model showed a 15% improvement in early detection of MDD (p < 0.001, McNemar’s test).

In the 2-year follow-up of initially healthy participants, the model correctly identified 78% (95% CI: 71%-85%) of individuals who later developed clinically diagnosed MDD.

### Interpretability

Activation maximization for the DNN model produced patterns consistent with reduced functional connectivity in the default mode network and hyperconnectivity in the salience network, aligning with current neurobiological theories of MDD.

These results suggest that our AI models, particularly the Deep Neural Network, show promising performance in detecting early-stage MDD using fMRI data. The models demonstrate good generalizability across different datasets and potential clinical utility in early identification of at-risk individuals. The identified important features align well with existing neurobiological understanding of MDD, providing a level of interpretability to the AI-driven approach.

## Discussion of fMRI-based AI Detection of Early MDD

Our study demonstrates the potential of machine learning models, particularly deep neural networks (DNNs), in detecting early-stage Major Depressive Disorder (MDD) using fMRI data. The high performance of our models, with the DNN achieving 89% accuracy and an AUC-ROC of 0.95, suggests that AI-driven analysis of neuroimaging data could serve as a valuable tool in the early diagnosis and risk assessment of MDD.

### Comparison with Existing Literature

Our findings align with and extend previous research in this field. For instance, Kambeitz et al. (2017)^17^ reported an AUC of 0.87 in their meta-analysis of machine learning models for MDD classification. Our superior performance (AUC 0.95) may be attributed to our use of more advanced algorithms and a larger, more diverse dataset. Moreover, our study’s focus on early-stage MDD represents a significant advancement, as most previous works have focused on already-diagnosed cases^18^.

### Neurobiological Insights

The importance of functional connectivity between the dorsolateral prefrontal cortex, anterior cingulate cortex, and limbic regions in our models is consistent with the neurobiological model of MDD proposed by Mayberg et al. (2005)^19^. These findings support the theory of disrupted emotional regulation circuits in MDD and suggest that these disruptions may be detectable in early stages of the disorder.

Our SHAP analysis highlights the reduced activation in the left dorsolateral prefrontal cortex during task-based fMRI. This corroborates previous findings by Koenigs and Grafman (2009)^20^, linking this region to cognitive control and emotion regulation deficits in MDD.

### Clinical Implications

The superior performance of our AI model compared to traditional DSM-5 criteria in early detection of MDD (15% improvement, p < 0.001) underscores the potential of this approach as an adjunctive tool in clinical practice. The model’s ability to identify 78% of individuals who later developed MDD suggests its potential use in preventive interventions.

However, it’s crucial to note that while our model shows promise, it should not replace clinical judgment but rather augment it. Integrating AI-based tools into psychiatric practice requires careful consideration of ethical implications and potential biases^21^.

### Limitations

Despite the promising results, our study has several limitations:

1. While our dataset was large and diverse, it may not fully represent all populations, potentially limiting generalizability.
2. The slightly lower performance in older participants warrants further investigation into age-related factors affecting model performance.
3. While informative, the 2-year follow-up period for assessing predictive capability may not capture very long-term outcomes.
4. Despite our efforts with techniques like SHAP, the interpretability of deep learning models remains a challenge.

### Future Directions

Future research should focus on:

1. Incorporating longitudinal fMRI data to capture the trajectory of brain changes in MDD development.
2. Integrating other data modalities (e.g., genetic and environmental factors) improves model performance and provides a more comprehensive risk assessment.
3. Conducting prospective clinical trials to validate the utility of these models in real-world clinical settings.
4. Developing more advanced interpretability techniques to enhance the clinical applicability of complex AI models.
5. Investigating the potential of these models in differentiating MDD from other psychiatric disorders with similar presentations.

## Conclusion

This study demonstrates the promising potential of artificial intelligence, particularly deep neural networks, in the early detection of Major Depressive Disorder (MDD) using fMRI data. Our findings reveal several key insights:

1. AI models, especially the deep neural network, achieved high accuracy (89%) and AUC-ROC (0.95) in detecting early-stage MDD, outperforming traditional diagnostic methods.
2. The models identified crucial functional connectivity patterns, particularly involving the dorsolateral prefrontal cortex, anterior cingulate cortex, and limbic regions, aligning with current neurobiological theories of MDD.
3. The AI approach demonstrated good generalizability across different datasets and showed promise in identifying individuals at high risk of developing MDD in a 2-year follow-up.
4. While powerful, these AI tools should be viewed as complementary to clinical judgment rather than replacements, with careful consideration given to ethical implications and potential biases.
5. Future research should focus on longitudinal studies, integrating multiple data modalities, and further enhancing model interpretability to bridge the gap between AI-driven insights and clinical application.

In conclusion, this study represents a step forward in leveraging AI for early detection of MDD. By enabling earlier and more accurate identification of at-risk individuals, this approach has the potential to transform clinical practice, allowing for more timely interventions and personalized treatment strategies. As we continue to refine these methods and address current limitations, the integration of AI-driven neuroimaging analysis into psychiatric care could play a crucial role in improving outcomes for individuals at risk of MDD.

## Data Availability

All data produced in the present study are available upon reasonable request to the authors.

## References

1. World Health Organization. (2021). Depression. https://www.who.int/news-room/fact-sheets/detail/depression

2. American Psychiatric Association. (2013). Diagnostic and statistical manual of mental disorders (5th ed.). Arlington, VA: American Psychiatric Publishing.

3. Patel, M. J., Khalaf, A., & Aizenstein, H. J. (2016). Studying depression using imaging and machine learning methods. NeuroImage: Clinical, 10, 115–123.

4. Wise T, Radua J, Via E, Cardoner N, Abe O, Adams TM, Amico F, Cheng Y, Cole JH, de Azevedo Marques Périco C, Dickstein DP. Common and distinct patterns of grey-matter volume alteration in major depression and bipolar disorder: evidence from voxel-based meta-analysis. Molecular psychiatry. 2017 Oct;22(10):1455–63.

5. Drevets, W. C., Price, J. L., & Furey, M. L. (2008). Brain structural and functional abnormalities in mood disorders: implications for neurocircuitry models of depression. Brain structure and function, 213(1-2), 93–118.

6. Mulders, P. C., van Eijndhoven, P. F., Schene, A. H., Beckmann, C. F., & Tendolkaro, I. (2015). Resting-state functional connectivity in major depressive disorder: A review. Neuroscience & Biobehavioral Reviews, 56, 330–344.

7. Lundervold, A. S., & Lundervold, A. (2019). An overview of deep learning in medical imaging focusing on MRI. Zeitschrift für Medizinische Physik, 29(2), 102–127.

8. Esteva A, Robicquet A, Ramsundar B, Kuleshov V, DePristo M, Chou K, Cui C, Corrado G, Thrun S, Dean J. A guide to deep learning in healthcare. Nature medicine. 2019 Jan;25(1):24–9.

9. Gao, S., Calhoun, V. D., & Sui, J. (2018). Machine learning in major depression: From classification to treatment outcome prediction. CNS neuroscience & therapeutics, 24(11), 1037–1052.

10. Kambeitz J, Cabral C, Sacchet MD, Gotlib IH, Zahn R, Serpa MH, Walter M, Falkai P, Koutsouleris N. Detecting neuroimaging biomarkers for depression: a meta-analysis of multivariate pattern recognition studies. Biological psychiatry. 2017 Sep 1;82(5):330–8.

11. Woo, C. W., Chang, L. J., Lindquist, M. A., & Wager, T. D. (2017). Building better biomarkers: brain models in translational neuroimaging. Nature neuroscience, 20(3), 365–377.

12. Varoquaux, G., & Poldrack, R. A. (2019). Predictive models avoid excessive reductionism in cognitive neuroimaging. Current opinion in neurobiology, 55, 1–6.

13. Yan B, Xu X, Liu M, Zheng K, Liu J, Li J, Wei L, Zhang B, Lu H, Li B. Quantitative identification of major depression based on resting-state dynamic functional connectivity: a machine learning approach. Frontiers in neuroscience. 2020 Mar 27;14:191.

14. Mourão-Miranda J, Oliveira L, Ladouceur CD, Marquand A, Brammer M, Birmaher B, Axelson D, Phillips ML. Pattern recognition and functional neuroimaging help to discriminate healthy adolescents at risk for mood disorders from low risk adolescents. PloS one. 2012 Feb 15;7(2):e29482.

15. Bzdok, D., & Meyer-Lindenberg, A. (2018). Machine learning for precision psychiatry: opportunities and challenges. Biological Psychiatry: Cognitive Neuroscience and Neuroimaging, 3(3), 223–230.

16. Jenkinson M, Beckmann CF, Behrens TE, Woolrich MW, Smith SM. Fsl. Neuroimage. 2012 Aug 15;62(2):782–90.

17. Kambeitz, J., Cabral, C., Sacchet, M. D., Gotlib, I. H., Zahn, R., Serpa, M. H., … & Koutsouleris, N. (2017). Detecting neuroimaging biomarkers for depression: a meta-analysis of multivariate pattern recognition studies. Biological psychiatry, 82(5), 330–338.

18. Gao, S., Calhoun, V. D., & Sui, J. (2018). Machine learning in major depression: From classification to treatment outcome prediction. CNS neuroscience & therapeutics, 24(11), 1037–1052.

19. Mayberg HS, Lozano AM, Voon V, McNeely HE, Seminowicz D, Hamani C, Schwalb JM, Kennedy SH. Deep brain stimulation for treatment-resistant depression. Focus. 2008 Jan;6(1):143–54.

20. Koenigs, M., & Grafman, J. (2009). The functional neuroanatomy of depression: distinct roles for ventromedial and dorsolateral prefrontal cortex. Behavioural brain research, 201(2), 239–243.

21. Char, D. S., Shah, N. H., & Magnus, D. (2018). Implementing machine learning in health care—addressing ethical challenges. The New England journal of medicine, 378(11), 981.

